# Neutralizing antibody responses 10 months after mild and moderately-severe SARS-CoV-2 infection

**DOI:** 10.1101/2021.02.22.21252225

**Authors:** Puya Dehgani-Mobaraki, Asiya Kamber Zaidi, Annamaria Porreca, Alessandro Floridi, Emanuela Floridi

## Abstract

Improved understanding of immunity offered by the antibodies developed against SARS-CoV-2 is critical. Our study aimed at longitudinal analysis of presence and persistence of neutralizing antibodies over ten months in mild and moderately-severe COVID-19 recovered patients using two immunoassays.

**Article Summary Line:** Neutralizing IgG antibody persistency was demonstrated in 63.3% of the subjects (19 out of 30) at ten months post-infection with zero re-infections.

## Introduction

The year 2021 began with large-scale rollout of vaccines against COVID-19 and the preliminary results in terms of acquired immunity post-vaccination have shown good results with reduction in the rate of infections and deaths. *(1)* However, long-term immunity in recovered patients is still a dilemma. Studies have shown the persistency of humoral immune response ranging from three months to more than eight months. *(2,3)* We evaluated the antibody responses of 30 people, based in the Umbria region in Italy, post severe acute respiratory syndrome coronavirus 2 (SARS-CoV-2) infection, over 10 months through six sequential serological tests.

### The study

A monocentric pilot longitudinal observational study was conducted on 114 participants, who had tested positive by real-time quantitative polymerase chain reaction (RT-PCR) for SARS-CoV-2 in the period between 1-30 March 2020. Written informed consent was obtained by all participants for voluntary participation in this study.The demographic characteristics, blood groups, co-morbidities, clinical features, treatment undertaken, and dates pertaining to symptom onset and RT-PCR tests were recorded. Sequential serum samples were collected over a period of ten months through six serial blood tests at appropriate intervals for 30 out of the 114 subjects who attended all the follow-up visits.

The aim of this study was to investigate for presence and persistency of SARS-CoV-2 specific antibodies over ten months using two commercial immunoassays :The MAGLUMI® 2019-nCoV lgM/lgG chemiluminescent analytical system (CLIA) Assay *(4)* for the first five tests and MAGLUMI® SARS-CoV-2 S-RBD IgG Chemiluminescence immunoassay (CLIA) for the last test. *(5)* These immunoassays were granted Emergency Use Authorization by the US Food and Drug Administration. Measurements and interpretation of results were made according to the manufacturer’s instructions. The subjects were divided into two groups based on disease severity: mild and moderately-severe. *(6)* Results were reported as measured chemiluminescence values divided by the cut off (absorbance/cut-off, S/CO): S/CO>1 was defined as positive and S/CO≤1 as negative. We treated time as a factor and defined six different time points; T0-T5; first blood sample was collected two months post-infection, in the month of May 2020 (T0) and then, one month (T1), three months (T2), five months (T3), six months (T4) and eight months (T5) after T0. The Association Naso Sano ethical committee approved the study (ANS-2020/001).

### Statistical analysis

Descriptive statistics for the main characteristics of the participants were expressed as Median, [1st-3rd] quartile for the continuous variables and as absolute frequency (column percentage) for the categorical variables. CLIA positivity cut-off was set at >1.01. Bar graph was used to determine the percentage of IgM and IgG positivity at each time point. In addition, a sub-analysis was performed dividing the sample into two severity groups (Mild and Moderately-Severe). Chi-squared test was used to measure the association between the severity groups and the categorical variables. The Mann U Whitney test was used to assess the differences between groups for the continuous variables at each time point. Shapiro Wilks tested the normal distribution of the data. The Friedman test was applied to look for statistically significant differences over time within the two severity groups for both IgM and IgG. All tests were two-sided, and a level of statistical significance was set at p<0.05. All the statistical analyses were performed using R software environment for statistical computing and graphics version 3.5.2 (R Foundation for Statistical Computing, Vienna, Austria;https://www.R-project.org/)

### Results

The data from 30 participants who had tested positive for SARS-CoV-2 infection were analyzed. The descriptive statistics for the main characteristics of the study group are reported in Table 1. The IgM and IgG titres divided into Mild (n=17) and Moderately-severe groups (n=13), expressed as a median [1st – 3rd] are reported in Table 2. Indeed, the p-value results from the Mann U-Whitney test. The CLIA positivity limit was set at >1.01 and the percentage of IgM and IgG positive subjects were analyzed over time, expressed as Bar plots. Anti-S-RBD IgG was detected in 19 out of 30 participants (63.3%) at T5, ten months post-infection. [Figure1].

**Figure 1.**
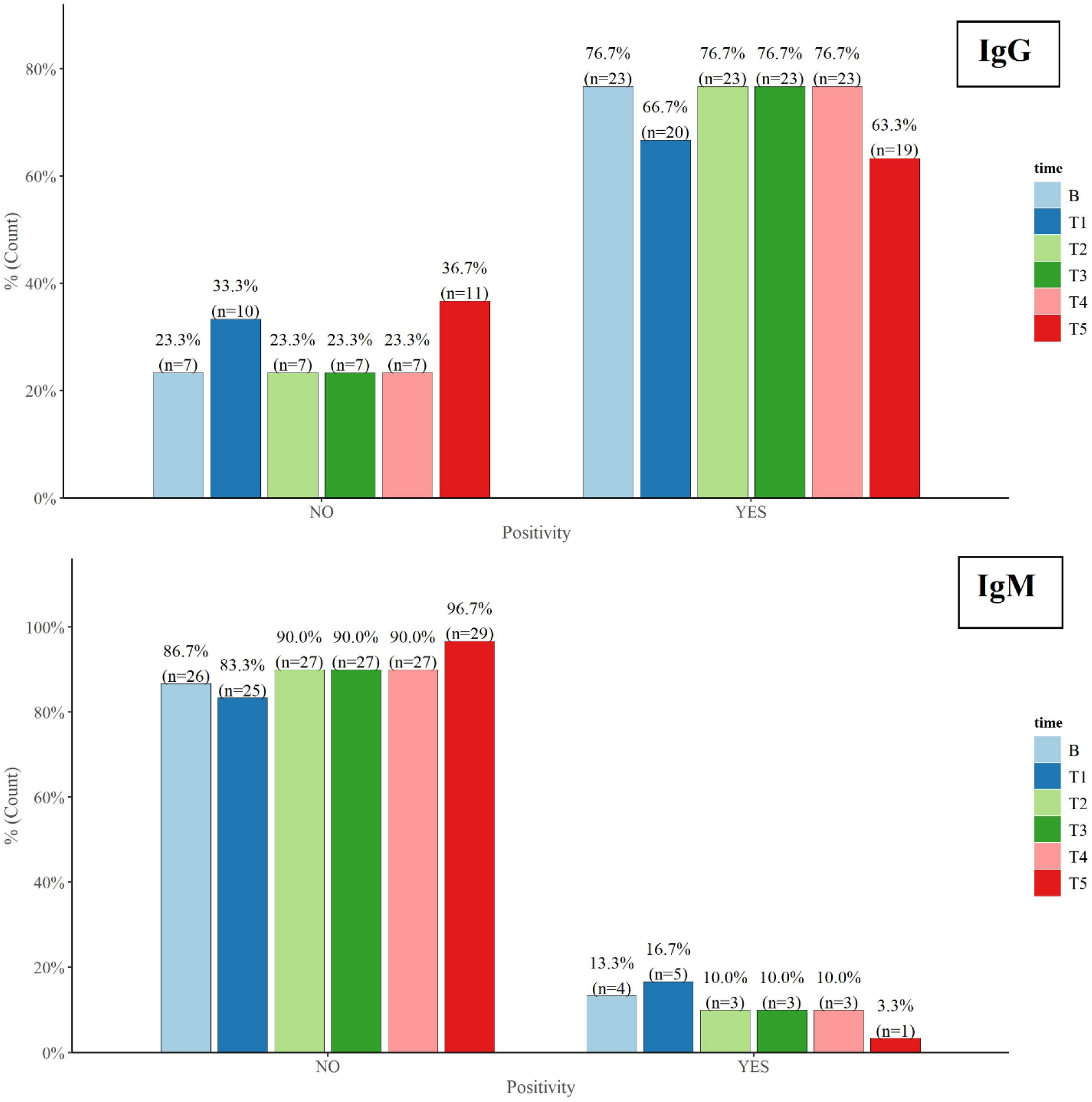
Bar plot for IgM and IgG positivity over ten months for all participants (n=30). (CLIA cut-off >1.10). B; Baseline, T0-T5 are the six sequential serological test time points.

**Table 1.**
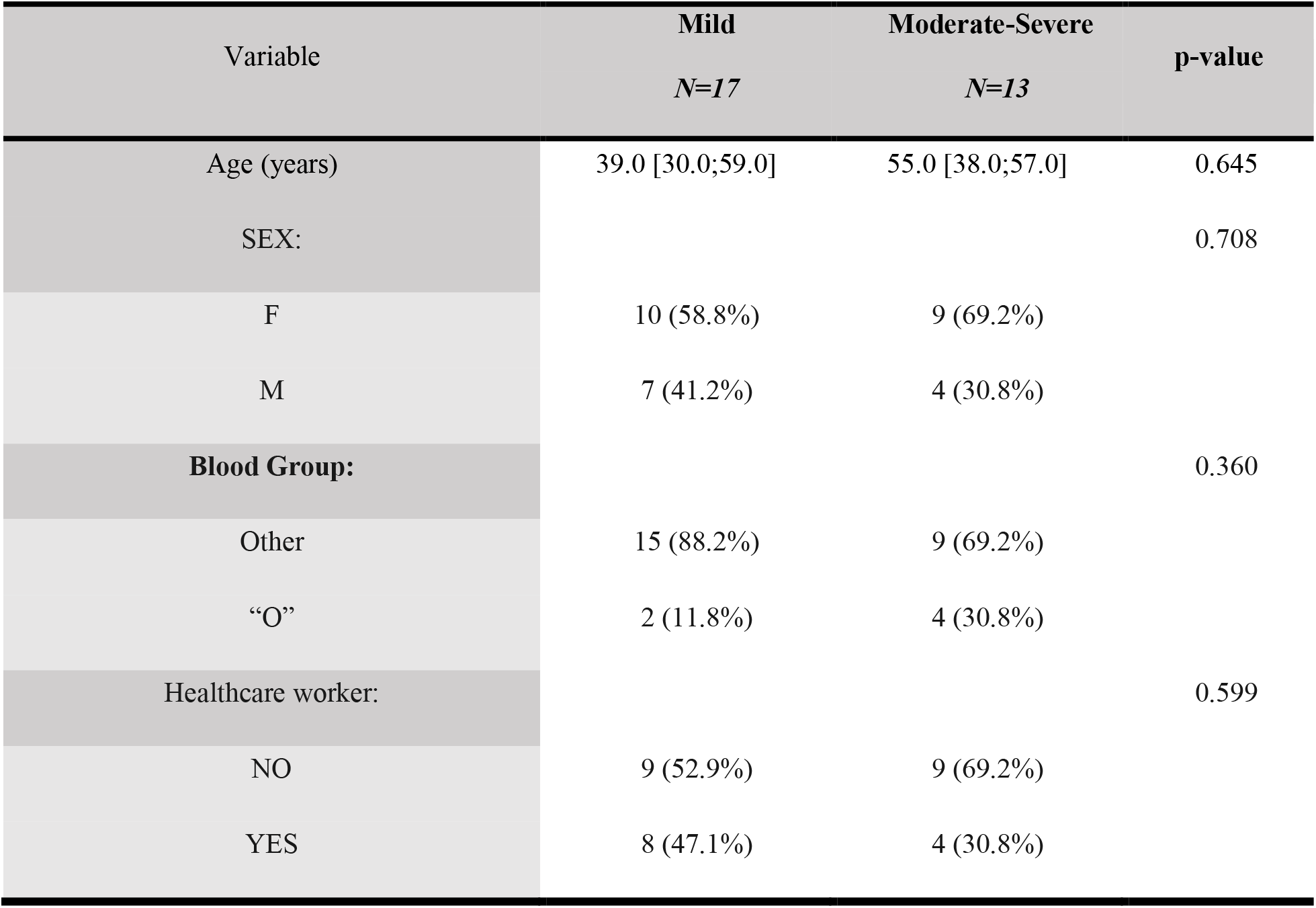
Descriptive statistics for the main characteristics of the subjects into the study (n=30) expressed as a median, [1^st^-3^rd^] quartile for Age and as absolute frequency (column percentage) for the categorical variables.

**Table 2.** The IgM and IgG titres, for the study group (n=30) divided into Mild (n=17) and Moderately-severe groups(n=13) expressed as a median [1st – 3rd]. The p-value results from the Mann U-Whitney test.

| Variable | Mild<br>*N=17* | Moderate-Severe<br>*N=13* | p-value |
| --- | --- | --- | --- |
| IgM.T0 | 0.50 [0.46;0.52] | 0.64 [0.64;1.07] | <0.001 |
| IgM.T1 | 0.61 [0.52;0.64] | 0.70 [0.66;1.44] | 0.006 |
| IgM.T2 | 0.36 [0.18;0.53] | 0.66 [0.24;0.84] | 0.194 |
| IgM.T3 | 0.23 [0.17;0.33] | 0.59 [0.31;0.82] | 0.030 |
| IgM.T4 | 0.21 [0.18;0.35] | 0.54 [0.25;0.81] | 0.057 |
| IgM.T5 | 0.03 [0.00;0.23] | 0.01 [0.00;0.16] | 0.897 |
| IgG.T0 | 1.84 [0.68;3.54] | 2.37 [1.14;20.7] | 0.621 |
| IgG.T1 | 1.81 [0.56;2.90] | 1.48 [0.83;20.6] | 0.832 |
| IgG.T2 | 1.70 [0.88;5.02] | 4.10 [2.38;16.1] | 0.269 |
| IgG.T3 | 3.05 [0.50;3.80] | 2.25 [1.04;10.2] | 0.724 |
| IgG.T4 | 3.13 [1.05;3.66] | 1.89 [0.83;8.53] | 0.724 |
| IgG.T5 | 1.46 [0.66;5.02] | 2.35 [0.32;5.06] | 0.868 |

The titres were higher for the female participants throughout the duration of 10 months (7, 5 and 2 females with 100%, 83.3% and 66.6% IgG positivity respectively Vs 7, 0 and 1 male with 100%, 83.3% and 66.6% IgG positivity respectively). Box plot diagrams for IgM and IgG for Mild (Panel 2A, Panel 2B) and Moderately-Severe (Panel 2C, Panel 2D) groups respectively are reported in Figure 2. For both groups, the IgM titre trend stayed below the set cut-off throughout time and also within the groups, changed in a statistically significant way (p<0.001). The IgG titre trend for both groups stayed above the positivity cut-off over time but did not change in a statistically significant way. (Mild: p=0.310, Moderately-Severe: p=0.370).

**Figure 2.**
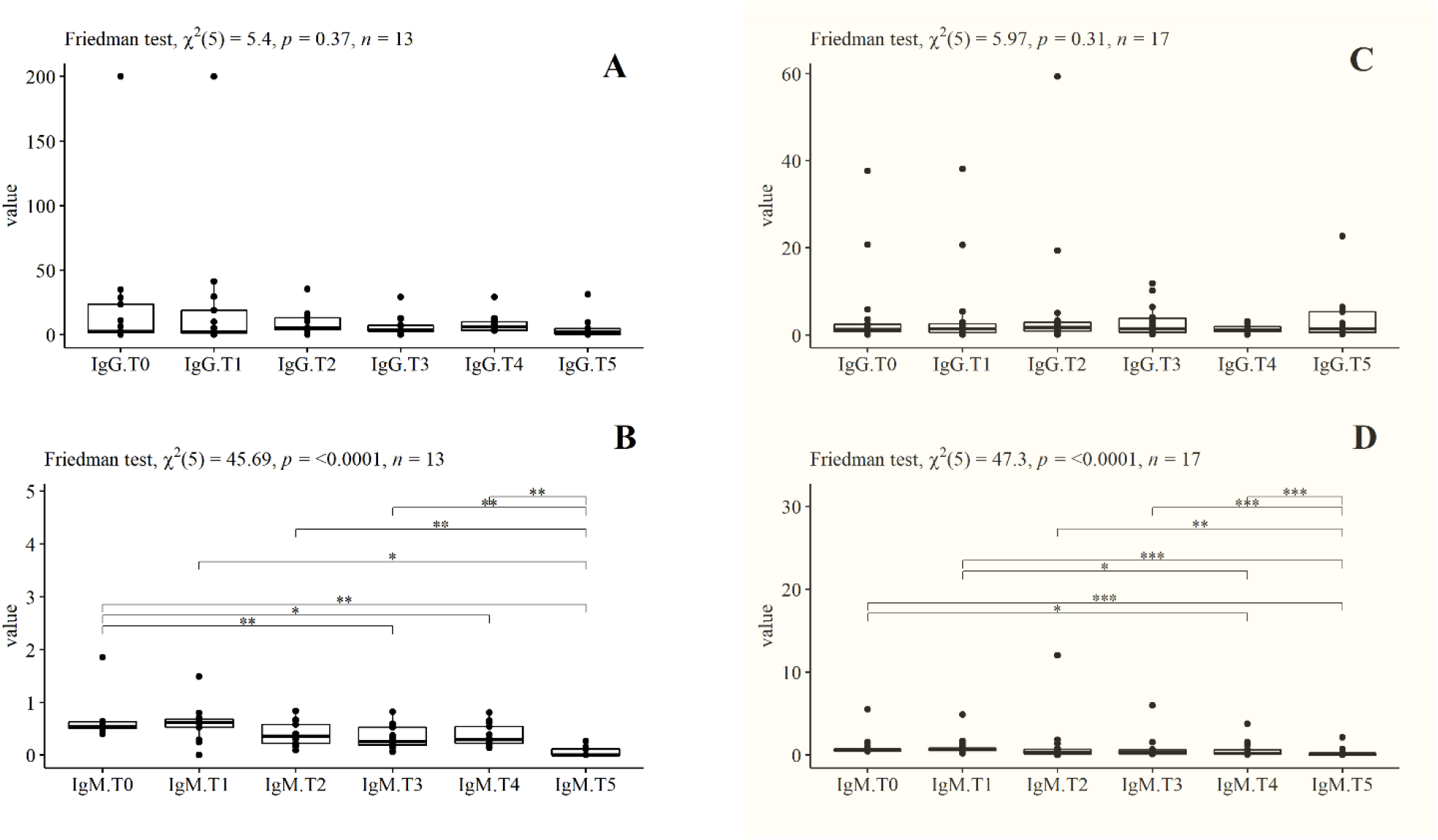
Friedman test for repeated measurements for IgG and IgM. Panel 2A and 2B are for the Moderately-Severe group (n=13) and Panel 2C and 2D are for the Mild group (n=17) (shaded as pale yellow). Pairwise comparisons have the following significance codes: 0 ‘***’ 0.001 ‘**’ 0.01 ‘*’ 0.05 ‘.’

### Discussion

SARS-CoV-2 is an enveloped beta-coronavirus with four structural proteins: Spike (S) protein, membrane (M) protein, envelope (E) protein and nucleocapsid (N) protein. Among the four proteins, the S protein and N protein are the main immunogens. The S protein is responsible for eliciting a highly potent neutralizing antibody response and is comprised of an N-terminal S1-subunit for virus-receptor binding and a C terminal S2-subunit for virus-cell membrane fusion. The S1-subunit is further divided into an N terminal domain (NTD) and a receptor-binding domain (RBD). This RBD directly interacts with host receptors to gain cell entry and is the most critical target for SARS-Cov-2 neutralizing Antibodies(nAb). *(7,8)* Antibodies against S protein can be detected six days post-PCR confirmation of infection, and those directed against the spike RBD show neutralizing capacity and could “prevent infection”. *(9,10)*

Recent study by Ibarrondo et al. have raised concerns regarding the duration of immunity offered by these antibodies. *(11)* Following this, Long et al. argued that “rapidly waning immunity” may lead to a substantial number of false-negative immunoassay results. *(8)* In contrast to these observations, some studies demonstrated the detection of antigen-specific memory T cells and memory B (Bmem) cells during convalescence. *(12,13)* Hartley et al. used unique sets of fluorescently-labeled recombinant tetramers of SARS-CoV-2 RBD and NCP antigens to “extensively characterize” the SARS-CoV-2-specific B mem cell response. The presence of circulating RBD- and NCP-specific Bmem cell subsets were detected early post-infection and persisted over 242 days post-symptom onset. These results also demonstrated that a decline in serum antibodies during the convalescent period might not reflect the waning of immunity, but rather a “contraction of the immune response” with “development and persistence of B cell memory”. *(14)*

An important point needs to be highlighted in terms of the association between humoral immune response and disease severity. A study by Choe et al. demonstrated that patients who might have experienced a relatively severe infection may develop an immunity that persists for a longer duration. *(3)* Similar results were observed in our study.

### Conclusion

The strengths of our study include diverse sample quality involving multiple family clusters of different age groups from the same region, adoption of CLIA as the method for analyzing the antibody titres and a long-term follow-up of 10 months post-infection. In a systematic review and meta-analysis by Bastos et al., pooled sensitivity with CLIA was 97.8% as compared to 84.3% with ELISA. *(15)* Our study reported zero cases of re-infection despite the fact that the Umbria region currently is experiencing a surge in daily cases with mutant strains.

## Data Availability

can be made available on request

## Acknowledgments

none

## References

1. Jeffay. N, 2021. It works: 0 deaths, only 4 severe cases among 523,000 fully vaccinated Israelis. [Online] Timesofisrael.com. Available at: <https://www.timesofisrael.com/hmo-sees-only-544-covid-infections-among-523000-fully-vaccinated-israelis/> [Accessed 19 February 2021].

2. Dehgani-Mobaraki. P, Kamber Zaidi. A, Floridi. A, Lepri. A, Floridi. E, Gherardi. A et al.. A comprehensive analysis of recovered COVID-19 patients and dynamic trend in antibodies over 3 months using ELISA and CLIA methods. medRxiv 2020.08.31.20184838; doi: https://doi.org/10.1101/2020.08.31.20184838

3. Choe PG, Kim K-H, Kang CK, Suh HJ, Kang E, Lee SY, et al. Antibody responses 8 months after asymptomatic or mild SARS-CoV-2 infection. Emerg Infect Dis. 2021 Mar [18 Feb 2021]. https://doi.org/10.3201/eid2703.204543

4. Drgmedtek.pl. 2021. MAGLUMI® 2019-nCoV lgG/lgM CLIA Assays. [Online] Available at: <https://drgmedtek.pl/wp-content/uploads/2020/02/M5001E01-MAGLUMI-2019-nCoV-IgG-IgM-200317.pdf> [Accessed 19 February 2021], MAGLUMI 2019-nCoV IgM/IgG - Letter of Authorization (fda.gov)

5. http://www.multimedilab.com/. 2021. MAGLUMI® SARS-CoV-2 Neutralizing Antibody (CLIA). x[Online] Available at: <http://multimedilab.com/wp-content/uploads/2021/01/MAGLUMI®-SARS-CoV-2-Neutralizing-Antibody-CLIA.pdf> [Accessed 19 February 2021].

6. https://www.who.int/publications/i/item/global-surveillance-for-human-infection-withnovel-coronavirus-(2019-ncov) WHO REFERENCE NUMBER: WHO/2019-nCoV/SurveillanceGuidance/2020.6

7. P. D. Burbelo, F. X. Riedo, C. Morishima, S. Rawlings, D. Smith, S. Das J.R. et al. Sensitivity in Detection of Antibodies to Nucleocapsid and Spike Proteins of Severe Acute Respiratory Syndrome Coronavirus 2 in Patients With Coronavirus Disease 2019. J. Infect. Dis. 222, 206–213 (2020). doi:10.1093/infdis/jiaa273pmid:32427334

8. Long QX, Tang XJ, Shi QL, Li Q, Deng HJ, Yuan J, et al. Clinical and immunological assessment of asymptomatic SARS-CoV-2 infections. Nat Med. 2020;26:1200–4.

9. M. S. Suthar, M. G. Zimmerman, R. C. Kauffman, G. Mantus, S. L. Linderman, W. H. Hudson, et al. Rapid Generation of Neutralizing Antibody Responses in COVID-19 Patients. Cell Rep Med 1, 100040 (2020). doi:10.1016/j.xcrm.2020.100040pmid:32835303

10. E. Seydoux, L. J. Homad, A. J. MacCamy, K. R. Parks, N. K. Hurlburt, M. F. Jennewein et al. Analysis of a SARS-CoV-2-Infected Individual Reveals Development of Potent Neutralizing Antibodies with Limited Somatic Mutation. Immunity 53, 98–105.e5 (2020). doi:10.1016/j.immuni.2020.06.001pmid:32561270

11. F. J. Ibarrondo, J. A. Fulcher, D. Goodman-Meza, J. Elliott, C. Hofmann, M. A. Hausner et al. Rapid Decay of Anti-SARS-CoV-2 Antibodies in Persons with Mild Covid-19. N. Engl. J. Med. 383, 1085–1087 (2020). doi:10.1056/NEJMc2025179pmid:32706954

12. J. A. Juno, H. X. Tan, W. S. Lee, A. Reynaldi, H. G. Kelly, K. Wragg et al. Humoral and circulating follicular helper T cell responses in recovered patients with COVID-19. Nat. Med. 26, 1428–1434 (2020). doi:10.1038/s41591-020-0995-0pmid:32661393

13. K. E. Lineburg, S. Srihari, M. Altaf, S. Swaminathan, A. Panikkar, J. Raju et al. Rapid detection of SARS-CoV-2-specific memory T-cell immunity in recovered COVID-19 cases. Clin. Transl. Immunology 9, e1219 (2020). doi:10.1002/cti2.1219pmid:33312565

14. Gemma E. Hartley, Emily S.J. Edwards, Pei M. Aui, Nirupama Varese, Stephanie Stojanovic, James Mcmahon et al. Rapid generation of durable B cell memory to SARS-CoV-2 spike and nucleocapsid proteins in COVID-19 and convalescence. Science Immunology 22 Dec 2020:vol. 5, Issue 54, eabf8891. DOI: 10.1126/sciimmunol.abf8891

15. Lisboa Bastos M, Tavaziva G, Abidi SK, Campbell JR, Haraoui LP, Johnston JC, et al. Diagnostic accuracy of serological tests for covid-19: systematic review and meta-analysis. BMJ. 2020;370:m2516

